# Perceptions, readiness and recommendations of traditional herbalists to integrate traditional and modern medicine in controlling COVID-19 epidemics in Northeast Ethiopia: An interpretive qualitative study

**DOI:** 10.1101/2022.12.01.22282963

**Authors:** Mesfin Wudu Kassaw, Mohammed Hussen Mohammed, Ousman Ahmed Mohammed

## Abstract

**Background:** Traditional medicine is an approach that has unique knowledge and beliefs which incorporates plant, animal or mineral based medicines that applied alone or in combination to treat, diagnose and prevent illnesses and maintain well-being. Suggestions from clinical practices and researches shown that integrated traditional Chinese and western medicine played an important role in China’s successful control of COVID-19. Despite such evidence, the Ethiopian minister of health prohibited traditional herbalists from using traditional remedies for COVID-19. However many of the traditional herbalists and the community requested the government frequently to try traditional medicine for COVID-19. The aim of this study was to explore perceptions, readiness, and recommendations of traditional herbalists on the effect of traditional medicine on COVID-19 and to select the promising remedies for pre-clinical study.

**Methods:** The study design used was an interpretive qualitative study. An in-depth interview was employed to gain access to the traditional herbalists’ experiences, perceptions, readiness and their recommendations. Traditional herbalists who lived in the North Wollo Zone were interviewed about the probable medicinal plants that can treat COVID-19. An inductive qualitative content analysis was conducted.

**Results:** From the in-depth interview with traditional herbalists, 4thematic frameworks were developed. Those major themes are;(1)perception of traditional medicine practitioners about COVID-19;(2) hypothesizing potential traditional remedies to treat COVID-19;(3)traditional practitioners recommendations for the community, and (4) integration of traditional and modern medicine. There was no pronounced difference in opinion among traditional herbalists about COVID-19 signs and symptoms, mode of transmission, and source of information about the epidemics.

Traditional herbalists had not planned to treat COVID-19 because of the minister of health’s prohibition of using traditional remedies. However, the traditional herbalists gave their remedies to minister of health, research institutes, and universities to get approval after the necessary procedures or laboratory investigations including toxicity studies. Despite the interest of traditional herbalists, currently, traditional medicine is not anymore economically and professionally useful for traditional herbalists because of many factors including the Ethiopian People Democracy Republic Front’s (EPDRF) government negative attitude, and its domination by the western medicine. Traditional herbalists were unsure which remedy might treat the COVID_19 but they believed that plants that were used to treat cough, acute respiratory distress syndrome (ARDS), and other respiratory infections might be used to control the signs and symptoms of COVID-19. If there is potential traditional remedy for COVID-19 from the traditional herbalists, integration of traditional medicine (TM) and modern medicine (MM) may be compulsory to manage COVID-19 effectively.

## Introduction

Traditional medicine is a health practices, approaches, knowledge and beliefs incorporating plant, animal and mineral based medicines, spiritual therapies, manual techniques and exercises, applied singularly or in combination to treat, diagnose and prevent illnesses and maintain well-being(1). Traditional medical practitioners (TMPs) in Ethiopia rely on an explanation of disease that draws on both the “mystical” and “natural” causes of illness and employ a holistic approach to treatment(2). Despite western medicine becoming more widespread in Ethiopia, Ethiopians tend to rely more on TM(3). There were not occasions that gave a great emphasis to traditional medicine just like the century of 2019 novel corona virus (COVID-19) incidence. In the late December 2019, a new type of corona virus was identified(4), and has been causing an increasing rate of pneumonia cases and deaths(5, 6). The WHO declared as a Public Health Emergency on 30January 2020. The diseases is characterized by flu-like symptoms including fever, cough, and severe acute respiratory distress syndrome(7, 8).

Studies reported that Chinese herbal formula such as San Wu Huangqin Decoction, Lianhuaqingwen Capsule, and Yinhuapinggan granule, possesses antiviral effects, which might be associated with blocking of the proliferation and replication of the viral particles, and that they might be able to improve lung damage by influenza viruses(9-11). According to WHO, there are no vaccines or antiviral treatments for COVID-19. However, China can control COVID-19 by integrating traditional Chinese and Western medicine (12-14). An evidence from clinical practice and research has shown that integrated traditional Chinese and Western medicine played an important role for China’s successful control of COVID-19(15). The Ethiopian minister of health prohibited traditional herbalists from using traditional medicine/healing to COVID-19. However, many of the traditional herbalists and community tried to force the government frequently to use traditional medicine. The aim of this study was to explore the perception, readiness, and recommendations of traditional herbalists on the effect of traditional medicine on COVID-19. The findings are expected to produce evidence about the probable impact of traditional medicine on COVID-19. The study is likely to be utilized by the government based on the recommendation. Stakeholders like health professionals, research institutes, and public universities will also utilize the findings for programming or educating the public.

### Aim and interview guidebook

#### Aim

The aim of this study is to explore perceptions, readiness and recommendation of traditional herbalists about medicinal plants to treat COVID-19 and its integration with modern medicine in Northeast Ethiopia. The semi-structured guidebook used to explore perceptions, readiness and recommendations of traditional herbalists about preferable medicinal plants for COVID-19 in the North Wollo Zone were based on the following 11questions. Traditional herbalists, and or religious leaders that live in North Wollo Zone were interviewed using those 11questions listed below and other probing questions.

1. What do you think about the newly identified disease-COVID-19?
2. Do you treat viral diseases like Hepatitis, influenza, measles, and others diseases before?
3. Do you treat diseases that have respiratory related signs like cough, ARDS, & fever?
4. Do you treat diseases that have diarrhea and vomiting?
5. How do you treat if the diseases cause frequent fatigue or tiredness?
6. Do you think that you can treat this disease-COVID-19?
7. What is your recommendation for Ethiopian regarding traditional medicine and COVID-19?
8. Can a patient get both traditional and modern medicine at a point for the diseases, and COVID-19?
9. If there is a system, do you agree to work in hospital/health center/health facility together with health care professionals?
10. If the system developed and allow working in health facilities, what requirements you need to be fulfilled?
11. Do you have a plan to treat COVID-19 patients?

## Methods

### Study setting, design, and period

The data were collected in North Wollo Zone, Northeast Ethiopia. The data collection period was from June 07/2020–July 05/2020. North Wollo is one of the administrative zones that located in Amhara Region. The region is located to the southwest of Ethiopia and has 15 administrative zones. The study were conducted on traditional herbalists or and religious leaders. This study used an interpretive qualitative study design (16, 17). The data analysis technique used was an inductive thematic framework(18).

### Sampling

In the North Wollo Zone, there were only 18 licensed traditional herbalists. The study participants were all of these 18 licensed traditional herbalists (purposive census sampling) and other 18 none-licensed traditional herbalists were selected using purposive sampling. However, the data were saturated on the 35^th^ of traditional herbalist to the pre-tested 11 semi-structured interview guidebook. Since all the traditional herbalists were requesting the government to treat COVID-19, no one opposed to participate in the study. The interview was take place in their garden or clinics to observe their capacity.

### Sampling and in-depth interview

A purposive sampling method was used to select traditional herbalists in the study area. Of the traditional herbalists and or religious leaders that lived in the North Wollo Zone, 35subjects were interviewed by face-to face platforms. The purpose of this study was to get an overall understanding about the significance of traditional medicine in treating COVID-19 epidemics during the first wave of COVID-19.

The interview was audio-taped and last one to two hours. The aim of the interview was to explore the perceptions, readiness and recommendations of traditional herbalists about preferable medicinal plants for COVID-19 infections.

In order to explore the perception of traditional herbalists’ perception further and to clarify their opinion, the following prompts were also used. “Can you elaborate about the plants that used to treat viral infections, or can you state the side effect of the plants you use for respiratory infections? Is there any point that you can add about COVID-19 and remedies for COVID-19, and etc.

### Data collection procedures and tools

The study populations were traditional herbalists in North Wollo Zone. A semi-structured guidebook was prepared based on the objectives and research questions of the protocol. The guidebook was designed in such a way with the assumption of requesting all traditional herbalists in similar fashion. The three researchers Mesfin Wudu Kassaw (BSc, MSc in Nursing), Mohammed Hussen Mohammed (BSc, MSc in chemistry), and Ousman Ahmed Mohammed (BSc, MSc in pharmacology) conducted the interview. All these researchers are male lecturers from Woldia Universities since most of the traditional herbalists were male. These three interviewers have a different work experience from 15years, 7years to 2years. The three interviewers conducted the study together to avoid bias and to facilitate a detail discussion with the traditional herbalists.

### Study participants

Traditional herbalists or and religious leaders who were working for more than five years on treating patients traditionally, and have license or planned to get license from national or region were selected for in-depth interview. The authors have no any relationship or contact with the traditional healers before the day of the interview. The authors described the purpose of the interview, their working place, and experience and gave to them a support letter from the district health department in which the traditional healers lived and from Woldia University. The traditional healers were had also a good understanding about the need for COVID-19 researches at that time.

### Quality assurance

In each step, standard operational procedures (SOP) were followed. A pretest was also conducted and the semi-structured interview questionnaires were edited accordingly for actual data collection. The interview guide developed in English, and translated to Amharic, and translated back to English. The major suspected bias in this study was religious-desirability bias. All the traditional herbalists were either Ethiopian Orthodox Tewahido Christian (EOTC) or Muslim, and the interviewers were 2Muslim and 1EOTC. To avoid bias, there was no anyone allowed to be there except the traditional healer and the three interviewers.

### Trustworthiness

Before the beginning of the study, a pretest was performed to ensure the validity and reliability of the semi-structured guidebook in Debre Tabore Town, Amhara Region, which far 50km from Bahir-Dar, the capital city of Amhara Region. Some of the ambiguities and unrealistic expressions in the semi-structured interview guidebook identified during the pre-test in Debre Tabore Town were corrected accordingly. The simplicity of the language utilized for interviews and descriptions to convey the findings were evaluated at all stages of the study. The interviews were taped and transcribed by three researchers. However, data coding had been done by two experienced and certified qualitative data analysts independently. The codes of the two qualitative researchers were similar and any discrepancy handled in discussion with the researchers. The audio record was narrated into text by two experienced translators independently. The difference between the audio record and narrated text were corrected through communication with interviewers and the study subjects involved in the interview and a further analysis was considered based on the suggestions.

The findings from the in-depth interviews were sent back to all the participants to confirm whether their concern was written correctly and to update the thematic frameworks. In addition, all the traditional herbalists were attending the dissemination presentation in Woldia University and forward their suggestions and the analysis was still updated. Thus, the validity of the findings was enhanced by employing study participants triangulation(19) and it was analyzed and re-analyzed frequently up to the end of the study. The credibility, dependability, conformability, transferability, and authenticity were considered ensuring trustworthiness(20).

To ensure credibility(16), we selected a maximum variation samples(licensed, non-licensed, religious fathers, ordinal fathers/mothers, male, female, Muslim, and Christian) to capture the range and variations of the first-hand perception of traditional herbalists(21-23). The interview had semi-structured open-ended questions, allowing the participants to speak freely, using their own stance(21). Data saturation was gotten as no new concepts emerged regarding the perceptions of traditional herbalists about medicinal plants regarding COVID-19. Dependability(16) was assured by conducting the interviews in a conversation-like manner to establish rapport, and unclear statements were asked to be clarified. The interviewer were trained to strove to listen attentively with an open mind(24). Field notes were taken immediately after each interview(17) when necessary. Thematic analysis was used to framework the interviewed texts(25). N-Vivo was used to group the transcribed verbatim by themes. Paragraphs were coded in N-Vivo for subsequent manual analysis of extracted material. Conformability(16) was assured by cross-checking the analysis with the guidebook and field notes(20). Transferability(16)were sought by providing detailed descriptions of all aspects of the study, helping readers to judge whether the findings were applicable in other contexts. To ensure authenticity(26), the findings reflect multiple realities and differences in functional ability at each phase of the trajectory.

### Data analysis

The transcripts were read many times to obtain a preliminary understanding of the participants’ experiences and the context(27). In the initial exploration of the interview, we observed that the participants described COVID-19 differently, and mentioned different type of medicinal plants. In addition, considerable differences were noted between the opinions of traditional herbalists. N-Vivo-10 software(28) were used to extract the data that were pertinent to the research questions, including sentences and text passages(29).

With the support of N-Vivo-10 software, an inductive thematic analytic approach was used (25, 30). Upon reading the extracted text, notes and open codes were written down in the margins(29). In the initial coding cycle(31), the meaning units were emerged freely from the text and were given a descriptive code (Table1).

We were revised the categories and sub-categories several times to ensure that the stated medicinal plants reflected the participants’ perceptions correctly(29).

**Table 1:**
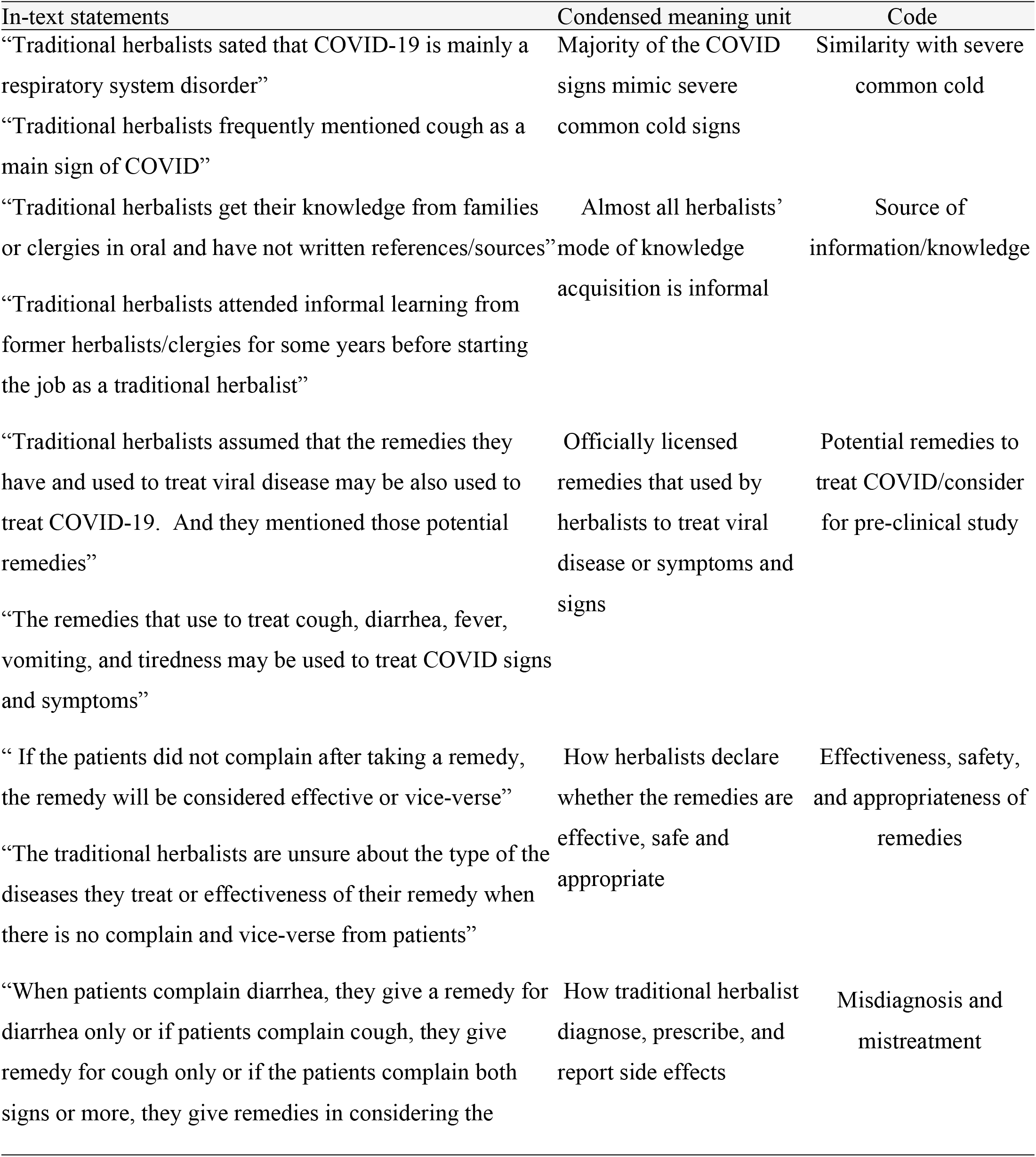
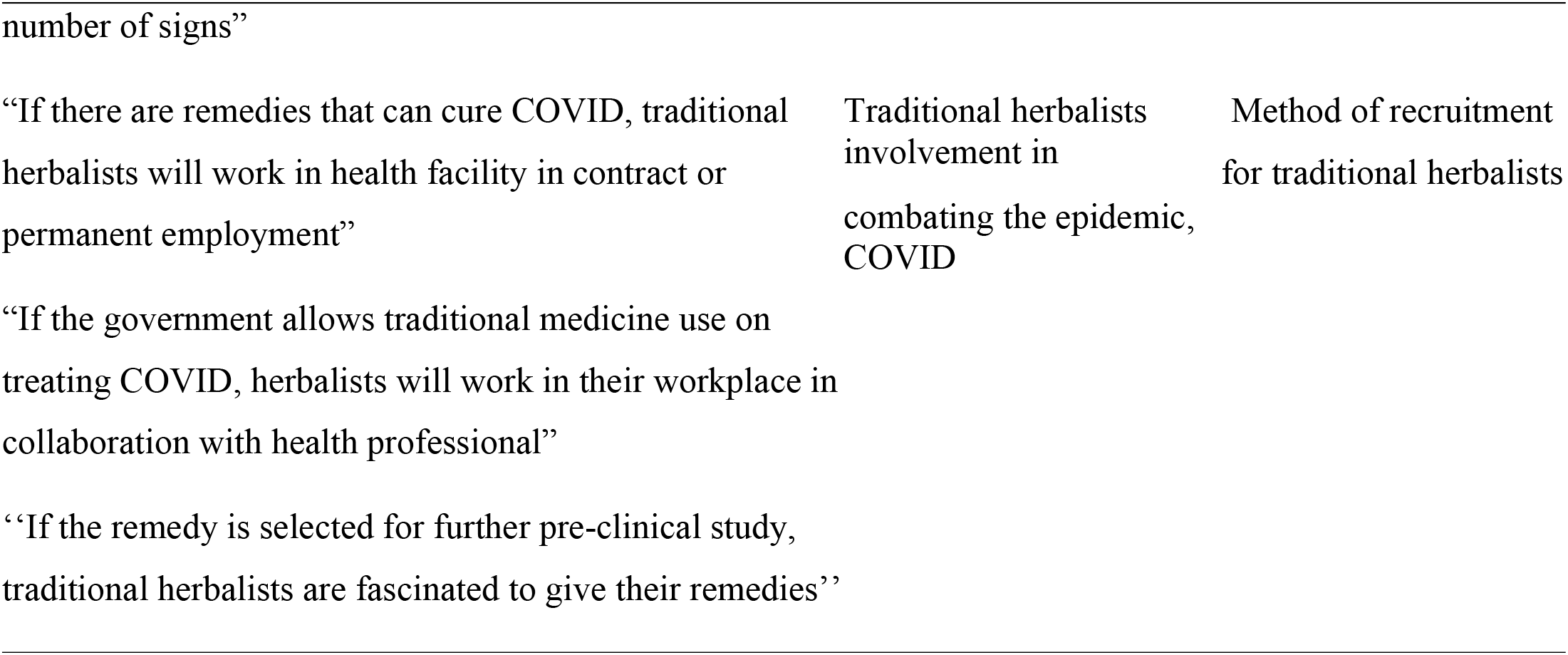
Examples of meaning units, condensed meaning units and codes.

### Terminologies

#### Perception

Traditional herbalist’s description of COVID-19 and its management from sensory impressions into a coherent and unified view

#### Readiness

Traditional herbalists’ preparation and availability of extracted medicinal plants or other healing options to treat COVID-19 in their home.

#### Recommendation

The traditional herbalists’ suggestion of potential medicinal plants to be used as a medicine for COVID-19.

#### Traditional Herbalists

A traditional practitioner that have no formal education but treating disease-using plants, animal and minerals only and or traditional healing like praying

#### Traditional Medicine

Any herbal, mineral, and animal remedies that could cure certain medical diseases

#### Herbal Medicine

plants that have a treating effect on certain human ailments and used by herbalists

#### Religious leaders

In this study, a religious leader is religious personnel who also practiced traditional healing using traditional medicine to cure patients

#### Ethics approval and consent to participate

An ethical clearance letter was obtained from Woldia University Institutional Review Board (IRB). A permission letter also collected from Woldia University Research and Development Director Office.

Before initiating the interview, oral informed consent obtained from traditional herbalists. The purpose of the study and its probable risks were clearly explained to the traditional herbalists. However, confidentiality of all information gathered were guaranteed, and used only for the stated purposes. When the traditional herbalists agreed, the name of the plant/animal/mineral that used to treat COVID-19 were requested and recorded. However, this question was optional since it had patent issue.

## Results

### Participant’s characteristics

The study subjects, traditional herbalists were living in 8 of the 15 north Wollo districts. The age of traditional herbalists ranged from 21 to 98 years old with a mean of 48.6(SD±13.99). Out of the 35study participants 32(91.4%) were males and 18(51.4%) of the traditional herbalists were licensed professional herbalists. Twenty one (60%) traditional herbalists got their knowledge through education (religious) and 13 (37.1%) were predominantly traditional herbalists who learned the knowledge by experience from their former relatives. Majority of the traditional herbalists were Orthodox United Christian in religion 19(54.3) and 8(22.9%) traditional herbalists were from Habru District (Table2).

**Table 2:** An example of category, sub-category and descriptive codes.

| Variables | Categories | Frequency | Percent (%) | Mean(Sd.) |
| --- | --- | --- | --- | --- |
| Gender | Male | 32 | 91.4 |  |
|  | Female | 3 | 8.6 |  |
|  | Total | 35 | 100 |  |
| Professional license | Licensed | 18 | 51.4 |  |
|  | Not licensed | 17 | 48.6 |  |
|  | Total | 35 | 100 |  |
| Profession | Orthodox united Christian Priest | 8 | 22.9 |  |
|  | Muslim clergy | 11 | 31.4 |  |
|  | Predominantly traditional healer | 13 | 37.1 |  |
|  | Others | 3 | 8.6 |  |
|  | (Teacher, Agricultural employee) |  |  |  |
| Mode of knowledge acquire | Family | 8 | 22.9 |  |
|  | Education (religious fathers) | 21 | 60.0 |  |
|  | Others (travelers to Sudan) | 6 | 17.1 |  |
| Age group | 20–29 | 1 | 2.9 | 48.60 |
|  |  |  |  | (13.99) |
|  | 30–39 | 8 | 22.9 |  |
|  | 40–49 | 13 | 37.1 |  |
|  | 50–59 | 7 | 20.0 |  |
|  | ≥60 | 6 | 17.1 |  |
|  | Total | 35 | 100 |  |
| Residence | Habru district | 8 | 22.9 |  |
|  | Woldia district | 6 | 17.1 |  |
|  | Meket district | 6 | 17.1 |  |
|  | Wadla district | 3 | 8.6 |  |
|  | Ayinabugna district | 6 | 17.1 |  |
|  | Lalibella district | 1 | 2.9 |  |
|  | Gidan district | 1 | 2.9 |  |
|  | Dawunt district | 4 | 11.4 |  |
| Religion | Orthodox united Christian | 19 | 54.3 |  |
|  | Muslim | 16 | 45.7 |  |

### Thematic frameworks

The data analysis of the study is based on thematic framework, and 4major were identified. Under the four major themes, a number of sub-themes were evolved further (Table3).

**Table 3:**
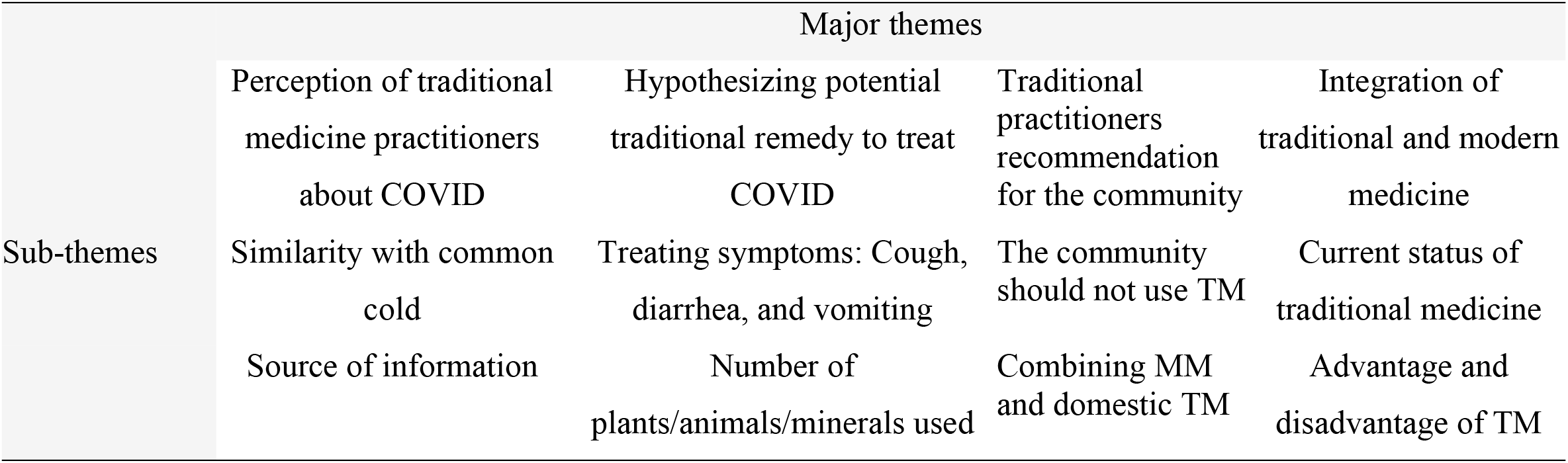
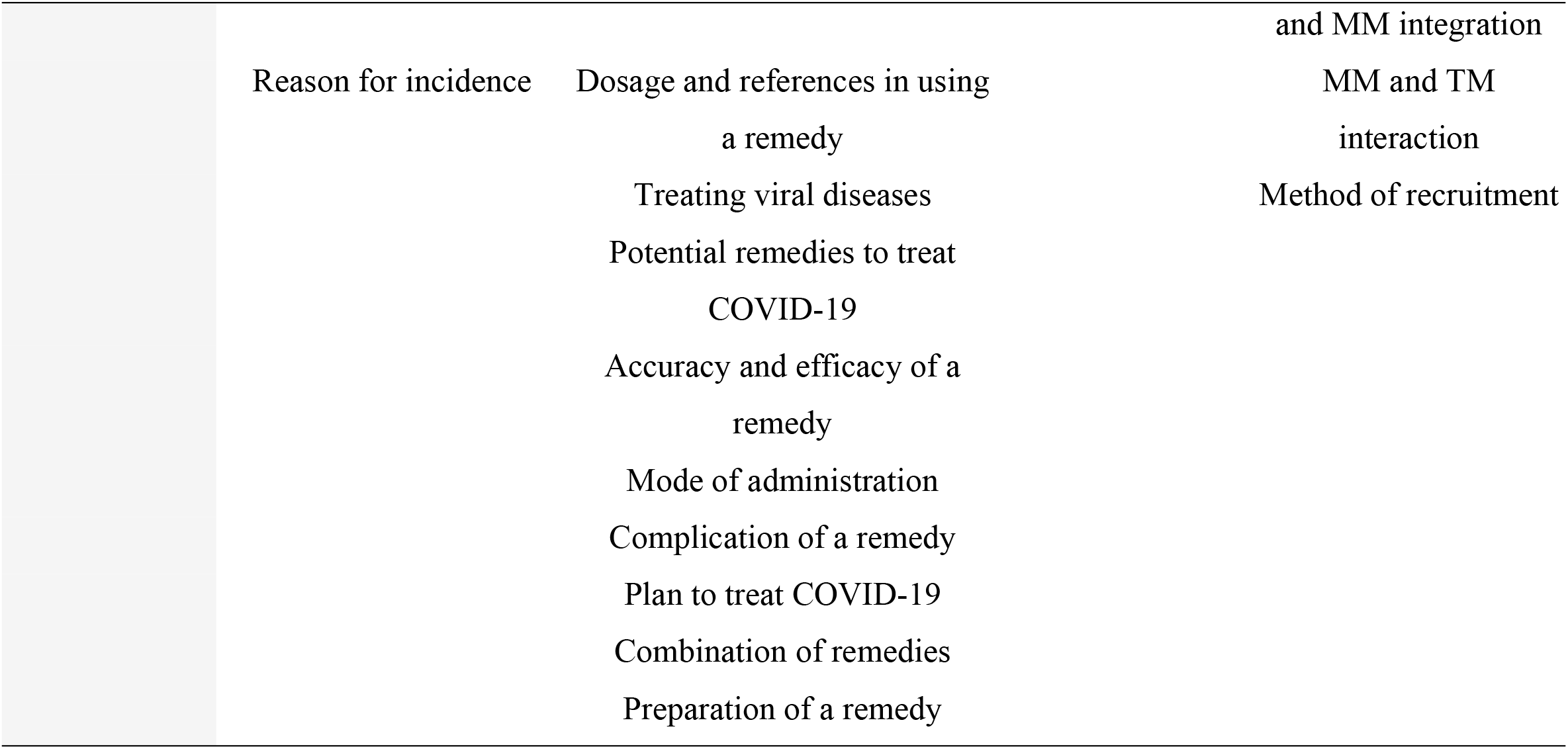
Socio-demographic characteristics of traditional herbalists in North Wollo Zone, Northeast Ethiopia.

### Theme I: Perceptions of traditional medicine practitioners about COVID-19

There was no great variability of opinion among traditional herbalists about COVID-19 signs and symptoms, mode of transmission, and source of information/knowledge. Almost all TMPs indicated that COVID-19 is a disease of respiratory tract. There was some variation on naming COVID-19. Most of them label it as l**ung locker**, and others define simply as it is a **lung disease**, and others mention it as **outbreak** of the 21^th^ century. In exploring perceptions of TMPs, three sub-themes were evolved further under ‘‘perceptions of traditional medicine practitioners about COVID-19’’ major theme as follow.

### Sub-theme-I: Similarity with common cold

The TMPs stated that COVID-19 is a type of common cold, and they believed that the symptoms of the disease can be treated using steam inhalation.

They perceived that COVID-19’s clinical features are exactly similar with severe common cold. One traditional herbalist described as; “the disease is more strong type of common cold and disseminate rapidly. This might be the difference with common cold” **TH15**

### Sub-theme-II: Source of COVID-19 information

The source of information they used to know about COVID-19 were locally placed health professionals, public Medias like television, and day to day communication within the community. Some traditional herbalists described the source of the information is legend or former prediction, because the signs and symptoms are exactly similar what they know by legend though they were not sure about the time of occurrence. But in legend, there was a prediction that affirmed the occurrence of common cold type diseases. They believed that COVID-19 is that and it had common cold feature which is exactly similar with the predicted diseases feature. One father stated that Satan was the other sources of information he used to know about the diseases, COVID-19. He usually calls the devil on a river and requests the unique events of the future, and then he got what he wants to know. “There was a legend that told to me before a long period of time about such worldwide pandemic disease, yet it was not called COVID-9. However, the signs and symptoms, the measurements to be taken and social impacts were already told to him”**TH16**. The researchers tried to affirm what he was saying by probing and observing his gesture, emotion, and confidence what he was talking about, however the topic was too sensitive and further description was not possible and the reflexivity of the researchers here was limited. One father described the legendary in relation to books as; “in addition to the oral legends, there are books (religious) that narrate the occurrences of epidemic in the 21^th^ century. Those religious books describe the details of the diseases including the prevention, controlling and managements options”**TH21**

### Sub-theme-III: Reason for COVID-19 incidence

The TMPs perceived that COVID-19 occurred because of God (super natural being) unhappiness with the current world view of people. They assumed that God is punishing us for our extreme impulsiveness. However, they believed that COVID-19 might not be happened in communities who practiced the recommended prevention measures like isolation, hand washing, or social distance. In addition, most TMPs accentuated more about the use of praying to clear-out the epidemics.

### Theme-2: Hypothesizing potential traditional remedy to treat COVID-19

There was a long lasting and deep two way interview based discussion with traditional herbalists about their remedies.

The traditional herbalists were interested to describe their remedies they used to treat viral diseases like influenza, HIV, measles, and herpes zosters on one side and remedies they used to manage symptoms of diseases like cough, diarrhea, and vomiting on the other side. They described the domestic and wild plants, animals as well as mineral sources of traditional remedies. In the hypothesizing potential traditional remedy to treat COVID-19 theme, 11sub-themes were evolved in developing the thematic framework during the analysis and described as follow.

### Sub-theme-I: Treating viral diseases

Some of the viral disease that TMPs treat includes hepatitis, skin symptoms of HIV, common cold, Herpes Zoster, measles, and mumps. Almost all TMPs used plants to treat viral diseases, but some of them also used animal products (mainly wild animals) to treat viral diseases. Some TMPs mentioned Bat as an example of animal source in treating viral diseases mainly hepatitis. But none of the TMPs use any mineral source in treating viral diseases. One traditional herbalist stated the importance of plants as; “plants are the foundations of human life and there are many types of plants that have many range of utility. For example there is a plant that called English (Amharic) In-vain (Etse-Bekentu) which has one day life span and used for many type of human ailments including viral diseases. I hope that this plant is needed in combination together with other plants in treating COVID-19” **TH8**

### Sub-theme-II: Treating COVID-19 symptoms: Cough, diarrhea, and vomiting

Traditional herbalists have different types of plants they used in preparing a remedy for those signs and symptoms. There are more than 16plants that TMPs used to treat cough. Sometimes, traditional herbalists used a single plant to treat both vomiting and diarrhea. They have also different plants for vomiting and diarrhea they used differently. A maximum of 5plants are used in treating vomiting and diarrhea.

### Sub-theme-III: Number of plants/animals/minerals used by TMPs

TMps mentioned that they used from 7plants (minimum) to 700plants (maximum) in treating their patients.

Of these ranges of plants used by the traditional herbalists, from 5(minimum) to 35(maximum) plants were used to treat viral diseases. They did not use minerals in treating diseases before but mentioned from 1 to 5minerals that can be used to treat human diseases.

Most of the TMPs are reluctant on the use of animals in treating diseases even they stated from 1 to 4animal source. This might be because of the Ethiopian culture in which almost all wild animals are not allowed to eat.

### Sub-theme-IV: Dosage and references in using a remedy

The main sources of knowledge for most TMPs were their family (usually their fathers), their former trainers, and Sudanese traditional herbalists. But as additional reference, they used books from bookstores, and religious articles like Koran in Muslim religion, and list and how to use medicinal plants book (Etse-Debdabie) in Ethiopian United Orthodox Christian religion.

### Sub-theme-V: Precision and efficacy of a remedy used by TMPs

Regarding the correctness, reliability and efficacy of their remedy, they have no scientific procedures they followed. However, they used their means of evaluation whether the remedy is safe, potent or not. The means they used is their complaints after using their remedy. If the patients visit them for repeated time after they took a specific remedy, they assume that the remedy is not effective for the sign they observe and reported from patient. Thus, they will not use it for the next time. Similarly, if their patients gave their appreciation for the new or old remedy regarding their chief complain, they record as the top remedy and used in another time for other patients. However, if they did not get evidence regarding its accuracy from their patients, they are unsure and used it as an adjunct until they got good testimonies or negative complain about it. “The sources we used are old books or old legends but the diseases are changing this day. Thus, the plants that worked before 50years may not work at this time and even the diseases may not present. Because of this, we are using different methods to increase our patients’ satisfaction from dose increment to combining different remedies” **TH9**

One priest also stated that “the only old reference book was Lists and how to use medicinal plants book (Este-Debdabie) from Ethiopia Orthodox Church might not be accurate 100% because the diseases are changing in relative to the reference book that were written before 1000years ago. Thus, we should update the remedy using patients complain”**TH12**

### Sub-theme-VI: Potential remedies to treat COVID-19

TMPs were unsure which remedy might treat the disease but they believed that plants that were used to treat cough, ARDS, and other respiratory tract problems might be used for COVID-19. Those all the proposed plants have been used to treat respiratory disorders predominantly. However, they are also mentioned some plants that was used to treat none respiratory problems. Most of the study participants were unwilling to tell the name of plants that would be used to treat COVID-19. They want a patent right or agreement with the organization that need the remedy. However, some of the TMPs stated Artemisia abyssinica **(Chikugn)** may be the best remedy on treating COVID-19 and preventing patients from having severe clinical signs and developing complications. One old traditional herbalist stated that “I am sure that **Self-pile/Horse-rod (Ras-Kimir)** can treat COVID-19 because I have been using it for different types of respiratory tract infections and has excellent efficacy”**TH11**

He has been administering orally with coffee for more than 30years and did not faced complain regarding this remedy. In addition to medicinal plants, TMP also mentioned many spice like domestic remedies that can be ingested with safe amount to treat COVID-19 in home as adjunct. Those domestic remedies to be ingested orally are (in Amharic) Ensilal, Azmerino, Zinjible, Garlic, Tena-adam, and Damakassie. Those domestic remedies have been used by the community as a spice or flavor in preparing meals. But the TMP also recommended inhaling Kerbie, and Etse-zeynone to treat and prevent COVID-19. Tilenji(Etse-hiwot), and Qotetina(Etse-debitera) were the other potential wild remedies to treat COVID-19 as mentioned by the traditional herbalists frequently. They also recommended Telenji ash in a home to prevent COVID-19. They also mentioned Gimero, Tembelel, Awulalet, and Ye enboay fire to be used together in treat COVID-19.

### Sub-theme-VII: Plan to treat COVID-19

TMPs had not planned to treat COVID-19 because of the minister of health’s prohibition on TMPs not to try to treat COVID-19 epidemics. However, they gave their remedies (medicinal plants) to Minister of health, research institutes, and public universities to get approval after the necessary procedures or laboratory investigations like toxicity study. They gave up to 7remedies that would be used in preventing or treating COVID-19. They were expecting that one of their remedy would have impact on preventing or treating COVID-19.

Nevertheless, they might give their remedy that has impact on COVID-19 and even for COVID-19 patients who visited them for other symptoms and got treatment because of COVID-19’s none-specific features. Because the remedies they have and gave to minister of health has been used to treat respiratory sign and symptoms including acute respiratory distress syndrome. They were waiting the invitation or permission from minister of health to treat COVID-19 locally or at centers. They also expect that the minister will provide trainings and develop appropriate precaution guidelines to prevent themselves from the disease as well from spreading in their community.

### Sub-theme-VIII: Mode of administrations

The mode of administration of the herbal remedies can be oral, topical, chewing, placing under a tongue, steam or inhalation for both domestic and wild types of remedies. TMPs perceived that those remedies can be used in both preventing and treating COVID-19 patients. The remedies used in the prevention of COVID-19 are steams that neutralize or kill the virus in a room if there are viruses confined there after occupied by COVID-19 positive patient. Most of the traditional herbalists stated to use a mixture of 7 and above plants to treat COVID-19 patients. Some TMPs explained about the way they would use to treat COVID-19. They believed that balm the whole body of the patient using the mixture of plants that soaked in water for 24hours would be the preferable option in treating COVID-19 patients. Because, most of TMPs perceived that the best remedies they have are topical that can be applied in the form of inhalation or ointment.

### Sub-theme-IX: Complications of remedies

TMPs appreciated that some of their remedies have well known side effects. But, they consider those side effects when administering or gave the remedies to their patients. However, many of the traditional remedies have not known whether they have side effects or not. Sometimes patients come with side effects after taking a remedy but many of patients might not visit the traditional herbalists even they develop complications.

### Sub-theme-X: Combination of remedies

TMPs usually prepare their remedies in single or in combination forms. They administer a single remedy (plant/animal/mineral) or a combination of different remedies for a number of signs and symptoms. They usually start with single or minimum number of combinations, but if the patient fails to have progress, they would add other remedies.

One traditional herbalist stated as; “the patient might complain up to 6 signs and symptoms. I would have 6 remedies for all of these 6 signs and symptoms but I do not give all the six remedies at the movement, rather I would give 3 for the most frequently happened signs or more severe signs. However, when the patient come again and complain, I will add the other remedies for each sign and symptoms step-wise **TH32**

### Sub-theme-XI: Preparation of remedies

TMPs prepare different remedies mainly composed of plant ingredients. Some of their preparations are single plant and other preparation contain up to 14plants. Almost all preparations have honey or others domestic remedies like Zinjible, Onion, Tena-adam, and Feto as adjunct.

### Theme III: Traditional practitioners’ recommendations

Traditional herbalists were asked about TM utilization for signs and symptoms of COVID-19 and their suggestion are framed in to two sub-themes as follow.

### Sub-theme-I: Avoid Traditional Medicine use

TMPs recommended avoiding TM utilization for the signs and symptoms of COVID-19 until the minister of health develop a guideline or update the regulations. They also perceived that the current habit of the community regarding the wild and domestic plant remedies was inappropriate. One herbalist described the habit of the community as; “most of the community ingest Onion or Feto even before knowing that they have COVID-19 or not. These remedies haven’t significant role in preventing COVID-19 but they may be used to treat COVID-19 to some extent. Nevertheless, these remedies have serious gastric side effects. Thus, the community should decide everything critically before using any of remedies even domestic remedies” **TH29**

### Sub-theme-II: Combining Western Medicine and Traditional Medicine

TM and MM can be taken together for a specific signs and symptoms but it does not work always. Some of the domestic remedies can be taken together with any medications. For example, Zinjible, Onion, green papper, Azmerino and etc. can be used together with a diet while the patient takes the usual medical care. But other remedies may have interaction with the modern medicine and cause serious complications.

### Theme-IV: Integration of traditional and modern medicine

If there is potential traditional remedy for COVID-19 that was used by traditional herbalists for other respiratory diseases, TM and MM integration may be compulsory for better effect. For this purpose, traditional herbalists were asked about the integration of TM and MM. They have raised a number of points for integration of TM and MM. But they stressed more about the platform how the minister of health and the traditional medicine association work together. To integrate TM and MM, four themes were evolved in the in-depth interview and described below.

### Sub-theme-I: Current status of traditional medicine

This day, traditional medicine is not useful for traditional herbalists because of many factors including the government, and domination of the western medicine. TMPs mentioned that traditional medicine is abolishing more than ever. Some of the plants are not get easily or everywhere or every time. Because, some plants can be gathered from deserts and others found in highlands. Some plants grow during summer and others grow during winter and due to such deviance, collecting remedies is really difficult but the price they received from patients is very low. One herbalists describes the challenge as; “when I was young, I collect up to 80plants from different parts of Ethiopia, but currently I only use the plants that are found in my garden and those are nearly only 50”**TH17**

### Sub-theme-II: Advantages and disadvantages of integrating traditional and modern medicine

Integrating TM and MM have its own advantages and disadvantages. The advantages of TM and MM integration includes; professional development for traditional medicine, increase patient satisfaction, increase the scope of clinical trial, professional satisfaction for traditional herbalists, and increase the efficacy of modern medicine. But it may have also cons like professional complexity, treatment errors, and professional malpractice.

### Sub-theme-III: Modern Medicine and Traditional Medicine interaction

TMP believed that some of the traditional medicine remedies may have interaction with modern medicine. But, they are not sure which of the remedy might have interaction with the modern medicines. Because of this lack of knowledge, traditional herbalists follow two precautions in administering their remedies. One, they are not give their remedy if the patient is taking modern medicine or have a plan to take prescribed or over the counter medicines. Second, they give their remedy after or before 1week of taking the prescribed medicine. But, they have also remedies that can be administer despite the patients’ modern medicine intake history.

### Sub-theme-IV: Methods of recruitment

TMPs have two points of view in working together with the health professionals under the Ethiopian health care system. The one is being recruited in a health facility as a health professional. The other is having traditional medicine dominated clinic that is supported by the government just like the health facilities that are governed by the government.

They also discussed the merits and de-merits of both view. If they are recruited in health facilities, they afraid that they may face a number of challenges like lack of plants. They also emphasized that they are familiar with traditional treatment cultures and may not be familiarize to the modern treatment culture easily. However, the second option is comfortable for both traditional herbalists and their patients. They raised points that the government should create a system for the traditional medicine just like the modern medicine. If there are well organized traditional medicine center, they can be also used as a research center. In addition, there may be created a referral communication between the modern and traditional health systems.

## Discussion

The purpose of this study was to assess the perception, readiness and recommendation of traditional herbalists to integrate traditional and modern medicine in treating COVID-19 in Northeast Ethiopia. The introduction of western medicine to Africa in the era of colonialism through Christian missionaries (32) led to the relegation of traditional medicine and the practitioners were derided and tagged ‘witch-doctors’ in some African countries like Nigeria (33) and Tenqoay in Ethiopia(34). Although this day traditional medicine is overlooked, both traditional and modern medicine were co-existing as two independent sectors, and respecting each other’s uniqueness and were had co-operation in different aspects such as mutual referral (35). Unlike evidence-based western medicine, traditional medicine is an empirical medicine developed on accumulated clinical observations gathered over centuries of practice. Traditional medicine is not only deals with the etiological factor to eradicate the pathogenic microbial, but also support the body’s immune function to help fight the disease and ameliorate its consequences. Contrasting to the Chinese traditional medicine that were used to treat SARS in which 58.3% of confirmed SARS cases received traditional Chinese herbal medicine(36), the Ethiopian traditional medicine had no role in controlling or treating outbreaks. Traditional herbalists recommended using potential traditional remedy together with western medicine to treat COVID-19. But, before using the remedies, at least toxicity study is needed, though traditional herbalists suggest a remedy that has been used for other diseases.

The recommendation of traditional herbalists agreed with a review (37) that found Chinese herbs combined with western medicine significantly improved symptoms of SARS, including decreasing body temperature, cough and breathing difficulties, and improve quality of life. However, another review of traditional Chinese herbal medicine for SARS revealed positive but inconclusive results about the efficacy of combining traditional Chinese herbal medicine and western medicine(38). Despite these evidences, the Ethiopian government does not invest on traditional medicine. Although, the government encouraged researches on traditional medicine, nothing is stated about the integration of traditional medicine and western medicine. However, the traditional herbalists recommended different remedies in different stages of diseases. Traditional herbalists told us that they gave up to 7remedies that would be used in preventing or treating COVID-19.

Herbalists were expecting that one of their remedy will have impact on preventing or treating the outbreak. But they were unsure about the effectiveness of their remedy in treating COVID-19. Yet they are expecting positive outcomes that their remedy will have impact because the remedies they have and gave to minister of health and research institutes has been used to treat respiratory signs and symptoms including acute respiratory distress syndrome previously. This is supported with a number of Chinese studies that reported that traditional Chinese herbal medicine was successfully prevented and treated SARS during the SARS epidemics (39-41). In addition, an evidence was mentioned about the importance of educating medical students and registered medical practitioners about TCAM therapies(42). Other evidence also stated that standardization; institutionalization and globalization of CTM are important issues for consideration in the setting of national and international public health research priorities, which includes operating principles that sufficiently fit the CTM in health care system(43). The integration needs to recruit traditional herbalists in a health facility. There is an evidence that entailed the importance of recruiting of traditional herbalists into a newly established scheme of community health care workers, and provide training to them for a new repertory of tasks(44).

Traditional herbalists are not sure which of the remedy ought to have interaction with the modern medicines. Because of this lack of knowledge, traditional herbalists follow two precautions in administering their remedies. They are not give their remedy if the patient is taking modern medicine or have a plan to take. Second, they give their remedy after or before 1 week of taking modern medicine. But evidences from China showed that combining traditional Chinese herbal medicine with western medicine regimen could reduce adverse events and other complications induced by Glucocorticoids, antibiotic, and antiviral treatments(45). Although, physicians have a lawful interest in their patients’ use of traditional remedies, particularly when there are known probable interactions with conventional medicine, up to 77% of patients do not disclose their use of CTM therapy to medical practitioners(46). This might be because of the traditional herbalists’ probation not to use modern medicine. Because of that most traditional herbalists did not know the side effects or the interaction with medicines. Hence, they prefer not to give their remedy or send to health facility if they want. Traditional herbalists recommended avoiding TM utilization for signs and symptoms of COVID-19 until the minister of health develop a guideline.

However, the Ethiopian government was not working well on forming expert team to formulate a traditional medicine treatment program unlike the Chinese government(36). Although, CTM was backed through a consumer or community supported movement in the past, it is slowly obtaining state support in the form of proactive national policies(47). But, the measurements taken by majority governments are not sufficient globally. But still, in this study, TMPs mentioned Zinjible, Garlic, green pepper, Tenadam, Azmerino and etc to be included with the western medicine. This is supported with a number of cases of cured patients from China who discharged from hospital after traditional Chinese herbal medicine treatment(7). Traditional herbalists describe that traditional medicine is not useful for traditional herbalists because of many factors including the government, and domination of western medicine. They mentioned that traditional medicine is abolishing more than ever. A study from Nigeria reported that traditional medicine is demoting back from health system and the practitioners were derided and tagged(33) especially after the western medicine introduced to the Africa continent in the wake of colonialism through missionaries of the Christian faith(32). However, evidence showed that the traditional medicine is growing and will lead to new research and business interests that may be ranged from providing affordable health care to developing new commercial products (43). This paradigm shifting is not coming true in Ethiopia rather a persisting negative attitude towards traditional medicine even by modern medical practitioners will pose uncertain future to ethno-therapy of the country(48). Traditional herbalists proposed plants to be used for COVID-19 that have been used to treat respiratory disorder signs with high degree of confidence. Most of the study participants were unwilling to tell the name of the plants that would be used to treat COVID-19. However, some of the participants mentioned that **Kopros (Chikugn) or Yeferes Zeng (Ras-Kimir)** will have an exceptional impact on COVID-19. This is supported by a retrospective study that have suggested Chinese medicine treat all mild and moderate COVID-19 cases and none became severe/critical after they took TCM (49).

## Conclusions

In the in-depth interview with the study participants, almost all traditional herbalists were sure about their remedies effectiveness on treating COVID-19. However, the minister of health’s probation hinders traditional herbalists from using their remedies. Most of the traditional herbalists have topical preparations that can be applied in the form of inhalation or ointment. However, most of the THs had not prescribed oral remedies and did not recommend for COVID-19 patients because of their inaccurate dose measurement and lack of experience which remedy may be preferable. But in conclusion, the government should involve actively in standardizing the extracts that will help in the stipulation of reliable information on the safety, efficacy and quality of herbal medicinal products.

## Limitations

Because of lack of researches conducted in Ethiopia; the discussion was supported by researches conducted abroad, mainly China. In relation to that, traditional medicine is going to disappear, there are no latest researches and we obeyed to cite obsolete articles. The other major limitation and future challenge will be the pray that traditional herbalists add when preparing or administering the remedy to their patient. However, the pray might be come from cultural practice as stated by Payyappallimana(47) and hope may have no relation with the efficacy of the plant. As indicated above the development of traditional medicine has been influenced by the different cultural and historic conditions. If the pray has an impact on the efficacy of the remedy, integration of traditional medicine and modern medicine will be difficult. Because the pray that traditional herbalists used vary from religion to religion and from society to society.

## Data Availability

All relevant data are within the manuscript and its Supporting Information files.

## Abbreviations

COVID-19: the 2019 novel corona virus
ARDS: Acute respiratory distress syndrome
TM: Traditional Medicine
MM: Modern medicine
THs: Traditional herbalists
TMPs: Traditional medical practitioners

## Declarations

### Ethics approval and consent to participate

An ethical clearance obtained from Woldia University institutional review board committee and it got a reference number of WDU/IRB-0076/2020.

An official permission letters were also collected from zonal health departments and town administrators. Written consent was obtained from all participants and when necessary the interviews were terminated at the behest of participants as well all the data was de-identified.

### Consent for publication

Not applicable

### Availability of data and material

The raw materials that support the conclusion of this research will be available to researchers needing the data to use for non-commercial purposes through requesting the authors through their e-mail.

### Authors’ contributions

For this study MWK, MHM, and OAM conceived the title and designed the study, preparing the data for analysis, analyzing the data, revising the first, second and final draft of this manuscript. All the authors read and approved the final version of this manuscript. The authors agreed to be accountable for all aspects of this work.

## Acknowledgment

We would like to thank Woldia University, college of health sciences, research and development office for providing full funding of this research. We would like to thank all the traditional herbalists for providing the required data. We are also acknowledging Woreda health office staffs, and zonal health department staffs for facilitating the communication with traditional herbalists.

## Reference

1. Zhang X. WHO Legal Status of Traditional Medicine and Complementary. Alternative Medicine: A Worldwide Review, WHO. 2001.

2. Bishaw M. Promoting traditional medicine in Ethiopia: a brief historical review of government policy. Social science & medicine. 1991;33(2):193–200.

3. Papadopoulos I, Lay M, Gebrehiwot A. Cultural snapshots: A guide to Ethiopian refugees for health care workers: Middlesex University; 2002.

4. World Health Organization. Novel coronavirus (2019-nCoV). Situation report. 2020;28.

5. Qun Li. An outbreak of NCIP (2019-nCoV) infection in China—wuhan, Hubei province, 2019− 2020. China CDC Weekly. 2020;2(5):79–80.

6. Tan W, Zhao X, Wang W, Niu P, et al. A novel coronavirus genome identified in a cluster of pneumonia cases—Wuhan, China 2019− 2020. China CDC Weekly. 2020;2(4):61–2.

7. Huang C, Wang Y, Ren L, Zhao J, et al. Clinical features of patients infected with 2019 novel coronavirus in Wuhan, China. The lancet. 2020;395(10223):497–506.

8. Chen N, Zhou M, Dong X, Gong F, Han Y, et al. Epidemiological and clinical characteristics of 99 cases of 2019 novel coronavirus pneumonia in Wuhan, China: a descriptive study. The Lancet. 2020;395(10223):507–13.

9. Xing X, Liu S, Shi C, Luo J. San wu. Huangqin decoction, a chinese herbal formula, inhibits influenza a/PR/8/34 (H1N1) virus infection in vitro and in vivo. Viruses. 2018;10(3):117.

10. Ding Y, Zeng L, Chen Q, Zhou B, Chen Q, et al. The Chinese prescription lianhuaqingwen capsule exerts anti-influenza activity through the inhibition of viral propagation and impacts immune function. BMC complementary and alternative medicine. 2017;17(1):130.

11. Zhou H, Yang J, et al. Antiviral effects and mechanisms of Yinhuapinggan granule against H1N1 influenza virus infection in RAW264. 7 cells. Inflammopharmacology. 2018;26(6):1455–67.

12. National Health Commission. Diagnosis and treatment protocol for novel coronavirus pneumonia (Trial Version 7). Chin Med J (Engl). 2020;133(9):1087–95.

13. Cheng S, Cheng H, Deng T, Fan P, et al. A rapid advice guideline for the diagnosis and treatment of 2019 novel coronavirus (2019-nCoV) infected pneumonia (standard version). Military Medical Research. 2020;7(1):4.

14. Deng Q, Peng J. Characteristics of and public health responses to the coronavirus disease 2019 outbreak in China. Journal of clinical medicine. 2020;9(2):575.

15. Qing G, Zhang H, Bai Y, Luo Y. Traditional Chinese and Western Medicines Jointly Beat COVID-19 Pandemic. Chinese Journal of Integrative Medicine. 2020:1.

16. Bryman A. Quantity and quality in social research: Routledge; 2003.

17. Lincoln YS, Guba EG. Naturalistic inquiry. Newberry Park. CA: SAGE; 1985.

18. Polit DF, Beck CT. Nursing research: Generating and assessing evidence for nursing practice: Lippincott Williams & Wilkins; 2008.

19. Saldaña J. The coding manual for qualitative researchers: SAGE; 2015.

20. Rabasa A. Radical Islam in East Africa: Rand Corporation; 2009.

21. Elo S, Kääriäinen M, Kanste O, Pölkki T, Utriainen K, Kyngäs H. Qualitative content analysis: A focus on trustworthiness. SAGE open. 2014;4(1):2158244014522633.

22. Patton MQ. Qualitative research and evaluation methods. Thousand Oaks. Cal: SAGE Publications. 2002.

23. Conrad P. The experience of illness: recent and new directions. Research in the sociology of health care Vol 6: the experience and management of chronic illness. 1987:1–31.

24. Morse JM, Barrett M, Mayan M, Olson K, Spiers J. Verification strategies for establishing reliability and validity in qualitative research. International journal of qualitative methods. 2002;1(2):13–22.

25. Kyngäs H. The qualitative content analysis process. Journal of advanced nursing. 2008;62(1):107–15.

26. Kvale S, Brinkmann S. Interviews: Learning the craft of qualitative research interviewing: sage; 2009.

27. Hsieh HF, Shannon SE. Three approaches to qualitative content analysis. Qualitative health research. 2005;15(9):1277–88.

28. Burnard P. A method of analysing interview transcripts in qualitative research. Nurse education today. 1991;11(6):461–6.

29. NVivo Q. QSR international Pty ltd. Doncaster, Victoria, Australia. 2002.

30. Schreier M. Qualitative content analysis in practice: Sage publications; 2012.

31. Downe-Wamboldt B. Content analysis: method, applications, and issues. Health care for women international. 1992;13(3):313–21.

32. Mbiti JS. African religions & philosophy: Heinemann; 1990.

33. Adefolaju T. Traditional and orthodox medical systems in Nigeria: The imperative of a synthesis. American Journal of Health Research. 2014;2(4):118–24.

34. Finseth KA, Finseth F. Health and disease in rural Ethiopia. The Yale Journal of Biology and Medicine. 1975;48(2):105.

35. Yoder PS. Biomedical and ethnomedical practice in rural Zaire: contrasts and complements. Social Science & Medicine. 1982;16(21):1851–7.

36. Liu X, Guo L, Zhong D, Zhang Y, et al. Traditional Chinese herbal medicine for treating novel coronavirus (COVID-19) pneumonia: protocol for a systematic review and meta-analysis. Systematic reviews. 2020;9:1–6.

37. Liu X, Zhang M, He L. Chinese herbs combined with Western medicine for severe acute respiratory syndrome (SARS). Cochrane Database of Systematic Reviews. 2012(10).

38. Leung PC. The efficacy of Chinese medicine for SARS: a review of Chinese publications after the crisis. The American journal of Chinese medicine. 2007;35(04):575–81.

39. Liu D, Liang B, Huang L. Clinical observation on the preventive effect of kangdu bufei decoction on acute severe respiratory syndrome. Zhongguo Zhong xi yi jie he za zhi Zhongguo Zhongxiyi jiehe zazhi= Chinese journal of integrated traditional and Western medicine. 2004;24(8):685–8.

40. Tong X, Zhang Z, Duan J, Chen X, Hua C, et al. TCM treatment of infectious atypical pneumonia--a report of 16 cases. Journal of traditional Chinese medicine= Chung i tsa chih ying wen pan. 2004;24(4):266–9.

41. Liu B, Liang Z, Tong X, et al. Effect of Chinese herbal medicine on adrenal glucocorticoids in SARS. China Journal of Chinese Materia Medica. 2005;30(23):1874–7.

42. Brooks PM. Undergraduate teaching of complementary medicine. Medical journal of Australia. 2004;181(5):275.

43. Bodeker G, Kronenberg F. A public health agenda for traditional, complementary, and alternative medicine. American journal of public health. 2002;92(10):1582–91.

44. Pillsbury BL. Policy and evaluation perspectives on traditional health practitioners in national health care systems. Social science & medicine. 1982;16(21):1825–34.

45. Hai X. Clinical experience of SARS treatment in Guangdong province. Tianjin Journal of Traditional Chinese. 2003;20(3):24–5.

46. Bishop FL, Yardley L, Lewith GT. Treat or treatment: a qualitative study analyzing patients’ use of complementary and alternative medicine. American Journal of Public Health. 2008;98(9):1700–5.

47. Payyappallimana U. Role of traditional medicine in primary health care: an overview of perspectives and challenging. 2010.

48. Abebe D, Ayehu A. Medicinal plants and enigmatic health practices of Northern Ethiopia. 1993.

49. Yuan W, Chen L, Han C, Zhang H, Luan X, et al. Combating COVID-19 with integrated traditional Chinese and Western medicine in China. Acta Pharmaceutica Sinica B. 2020.

